# Transcriptomics Analyses of ALS Post-mortem Motor Cortex highlight alteration and potential biomarkers in the Neuropeptide Signalling pathway

**DOI:** 10.1101/2023.05.05.23289551

**Authors:** Renata Kabiljo, Heather Marriott, Guy P Hunt, Abigail L Pfaff, Ahmad Al Khleifat, Brett Adey, Ashley Jones, Claire Troakes, John P Quinn, Richard J B Dobson, Sulev Koks, Ammar Al-Chalabi, Alfredo Iacoangeli

**Author notes:** R.K. and H.M. have contributed equally to the manuscript.

## Abstract

**Background:** Amyotrophic lateral sclerosis (ALS) is a fatal heterogeneous neurodegenerative disease that typically leads to death from respiratory failure within two to five years. Despite the identification of several genetic risk factors, the biological processes involved in ALS pathogenesis remain poorly understood. The motor cortex is an ideal region to study dysregulated pathological processes in ALS as it is affected from the earliest stages of the disease. In this study, we investigated motor-cortex gene expression of cases and controls to gain new insight into the molecular footprint of ALS.

**Methods:** We performed a large case-control differential expression analysis of two independent post-mortem motor cortex bulk RNA-sequencing (RNAseq) datasets from the King’s College London BrainBank (N = 171) and TargetALS (N = 132). Differentially expressed genes from both datasets were subjected to gene and pathway enrichment analysis. Genes common to both datasets were also reviewed for their involvement with known mechanisms of ALS pathogenesis to identify potential candidate genes. Finally, we performed a correlation analysis of genes implicated in pathways enriched in both datasets with clinical outcomes such as the age of onset and survival.

**Results:** Differential expression analysis identified 2,290 and 402 differentially expressed genes in KCL BrainBank and TargetALS cases, respectively. Enrichment analysis revealed significant synapse-related processes in the KCL BrainBank dataset, while the TargetALS dataset carried an immune system-related signature. There were 44 differentially expressed genes which were common to both datasets, which represented previously recognised mechanisms of ALS pathogenesis, such as lipid metabolism, mitochondrial energy homeostasis and neurovascular unit dysfunction. Differentially expressed genes in both datasets were significantly enriched for the neuropeptide signalling pathway. By looking at the relationship between the expression of neuropeptides and their receptors with clinical measures, we found that in both datasets *NPBWR1, TAC3* and *SSTR1* correlated with age of onset, and *GNRH1, TACR1* with survival. We provide access to gene-level expression results to the broader research community through a publicly available web application (https://alsgeexplorer.er.kcl.ac.uk).

**Conclusion:** This study identified motor-cortex specific pathways altered in ALS patients, potential molecular targets for therapeutic disease intervention and a set of neuropeptides and receptors for investigation as potential biomarkers.

## Background

Amyotrophic lateral sclerosis (ALS) is a devastating, heterogeneous neurodegenerative disease that leads to death from neuromuscular respiratory failure within two to five years. Despite the heritability of ALS being largely unknown, the discovery of several genetic risk factors has driven the identification of multiple biological processes involved in the disease aetiology^1,2^. However, the pathogenic mechanisms of ALS remain poorly understood.

The motor cortex is affected from the earliest stages of the disease and is one of the brain regions majorly affected by ALS pathology. Consequently, its disease expression signatures have been investigated to gain insight into the disease aetiology. Recent studies have reported differential expression of genes related to transcription, neurotransmission, protein synthesis and oxidative stress processes^3^, with extensive RNA expression alterations of neuroinflammatory and synaptic-related pathways identified at both gene and isoform levels^4^. Nevertheless, studies of this critical region at the whole-transcriptome level usually involve relatively small sample sizes and do not replicate their results in independent datasets.

To study functional changes occurring in the motor cortex of ALS patients, we have utilised two large independent RNAseq datasets of ALS patients and controls from the King’s College London (KCL) BrainBank (171 subjects) and TargetALS (234 samples from 132 subjects). By analysing the two datasets, our goal was to obtain profiles of altered gene expression that would identify the underlying molecular footprint of ALS and use this information to propose novel hypotheses about disease mechanisms, potential drug targets and biomarkers.

## Methods

### Datasets

The KCL BrainBank RNAseq dataset^5,6^ consists of post-mortem RNAseq samples from KCL and the Medical Research Council (MRC) London Neurodegenerative Diseases Brain Bank. Frozen human post-mortem tissue was taken from the primary motor cortex. The RNAseq protocol is outlined in the Supplementary Methods. 112 sporadic ALS cases and 59 controls were included in this analysis.

The TargetALS RNAseq dataset comes from TargetALS Human Postmortem Tissue Core and the New York Genome Centre (NYGC). We obtained the TargetALS dataset from https://www.targetals.org/. A list of the sample IDs of 243 samples (from 132 subjects, 112 ALS cases and 20 controls) used in this study is available in *Supplementary Table 1*. Frozen human post-mortem tissue was taken from the motor cortex (medial, lateral, and unspecified). Samples from different motor cortex regions for the same subjects were included, where available (see Supplementary Methods: TargetALS Sample Selection for more information on the inclusion of multiple samples per subject). Library preparation and sequencing of RNA are described in detail here https://www.targetals.org/wp-content/uploads/2020/11/README-1.zip.

### Bioinformatics, data processing and expression analysis

Both datasets were processed using the same pipeline, which is documented and distributed here: https://github.com/rkabiljo/RNASeq_Genes_ERVs. The pipeline involves interleaving the paired reads using reformat.sh from BBtools^7^ and trimming adapters and low-quality reads using bbduk.sh^7^, before aligning to the Hg38 reference genome using STAR^8^.Transcripts are then quantified using HTSeq^9^. The Supplementary Methods provides more details about each one of these steps and the parameters used. DESeq2^10^ was used for normalisation and differential expression analysis. The scripts are available at https://github.com/rkabiljo/DifferentialExpression_Genes. To test whether the number of differentially expressed genes identified in our analysis was due to chance, we performed 1,000 random permutations of case/control labels and performed differential expression analysis on each permutation. For each permutation, we counted the number of genes with false discovery rate (FDR) adjusted p-value lower than our selected cut-off of 0.05. The random permutation scripts are available here https://github.com/rkabiljo/ShufflingLabelsForDE. Functional enrichment was performed using MetaCore™ (available at https://portal.genego.com) for Gene Ontology (GO) processes, Pathway Maps, Process Networks and Disease by Biomarker enrichments, whilst gProfiler2^11^ was used for Reactome^12^ enrichment. We used BRETIGEA^13^ to derive cell proportions for the following cell types; astrocytes, endothelial cells, microglia, neurons, oligodendrocytes, and oligodendrocyte progenitor cells (OPCs).

To share the results with the wider community, we developed a web application using the R “shiny” framework (version 1.7.1) to allow quick visualisation of specific gene expression in ALS cases and controls, available at https://alsgeexplorer.er.kcl.ac.uk.

## Results

### Sample Characteristics

The summary characteristics of the two datasets are shown in Table 1. The KCL BrainBank dataset had significant differences between cases and controls in terms of age, RNA integrity number (RIN) and post-mortem delay (PMD). The TargetALS dataset showed no significant differences in age, sex, and RIN between cases and controls, but did display a significant difference in PMD (p-value = 0.0086). We have accounted for these differences through our choice of covariates included in the differential expression analyses (see Supplementary Methods). Principal component analysis (PCA) plots of samples based on expression data using the 500 most variable genes are in Supplementary Figures *1A* (KCL BrainBank) and *1B* (TargetALS).

**Table 1.** KCL BrainBank and TargetALS sample characteristics. Age: Age at death. PMD: Post-mortem delay. RIN: RNA integrity number. *p-value of Chi-square test for categorical variables (Sex) and T-test for continuous variables (Age, RIN Number, PMD)

|  | KCL BrainBank |  | *p*-value* |
| --- | --- | --- | --- |
|  | ALS Cases | Controls |  |
| **Total** | 112 | 59 |  |
| **Age (mean ± SD)** | 68.81±12.71 | 76.19±14.58 | 1.76e-06 |
| **Sex** |  |  |  |
| **Female (%)** | 47 (41.96%) | 33 (55.93%) | 0.11 |
| **Male (%)** | 65 (58.04%) | 26 (44.07%) |  |
| **RIN Number (mean ± SD)** | 6.44±1.26 | 5.33±1.58 | 8.38e-06 |
| **PMD (mean ± SD)** | 26.05±12.08 | 37.56±19.34 | 7.61e-05 |
|  | Target ALS |  | *p*-value* |
|  | ALS Cases | Controls |  |
| **Total** | 112 | 20 |  |
| **Age (mean ±SD)** | 63.95±10.57 | 62.6±11.86 | 0.64 |
| **Sex** |  |  |  |
| **Female** | 48(42.86%) | 11(55%) | 0.45 |
| **Male** | 64 (57.14%) | 9 (45%) |  |
| **Sample Characteristics** |  |  |  |
| **Total** | 198 | 36 |  |
| **RIN Number (mean ± SD)** | 6.20±1.48 | 5.91±1.25 | 0.2 |
| **PMD (mean ± SD)** | 10.49±6.92 | 21.16±22.84 | 0.0086 |

### Differential expression analysis identifies genes linked to ALS pathogenesis shared in both datasets

#### KCL BrainBank

A total of 2,290 genes were differentially expressed with an adjusted *p*-value of <0.05, in the KCL BrainBank dataset. Of these, 1,507 of the genes were upregulated and 783 were downregulated in cases (*Figure 1A, 1B*). None of the 1000 random permutations produced more differentially expressed genes than the actual differential expression analysis, thus confirming the significance of these results. In the vast majority of the permutations, there were no differentially expressed genes (with adjusted p-value <0.05), with the number of significant genes across permutations averaging 4.5.

**Figure 1.**
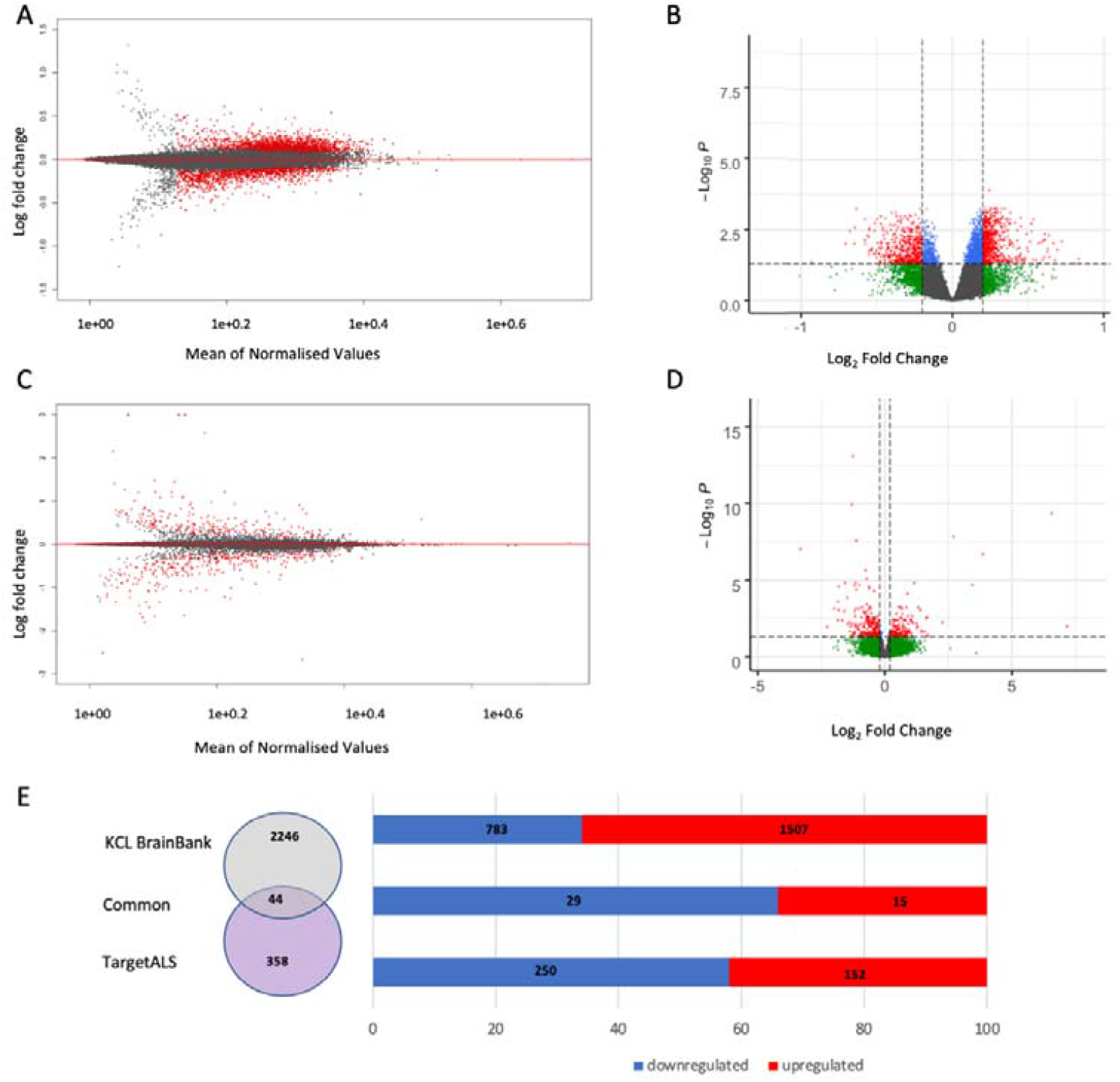
A) KCL BrainBank and C) TargetALS MA plots representing the distribution of log_2_ fold change (logFC) across all expression levels; B) KCL BrainBank and D) TargetALS Volcano plots. Plot D has been cropped to exclude one point with extremely high log_2_ fold change. Genes coloured red pass both the p-value and logFC thresholds. Grey genes are not significant, green genes only pass the logFC threshold, and blue genes only pass the logFC threshold. E) Numbers of differentially expressed genes in the two datasets.

#### TargetALS

A total of 402 genes were differentially expressed, with an adjusted p-value of <0.05, in the TargetALS dataset. Of these, 250 of these genes were downregulated and 152 were upregulated in cases (*Figure 1C, 1D*). When performing random permutations, 9 (0.9%) produced as many or more differentially expressed genes as the actual differential expression analysis. In most of the permutations, there were no differentially expressed genes (with adjusted p-value <0.05), with an average of 28 significantly expressed genes across permutations.

#### Shared Differentially Expressed Genes

We found 44 genes significant in both datasets with an adjusted *p*-value of <0.05. All of these genes had a concordant direction of regulation in both datasets; 29 genes were downregulated and 15 were upregulated (Figure 1E). To estimate the significance of this intersection between differentially expressed genes in the two datasets, we performed a hypergeometric test, using 41,709 as the number of background genes. The *p*-value obtained was < 1.2e-05. Genes that were significant in both datasets are listed in *Table 2*.

**Table 2.**
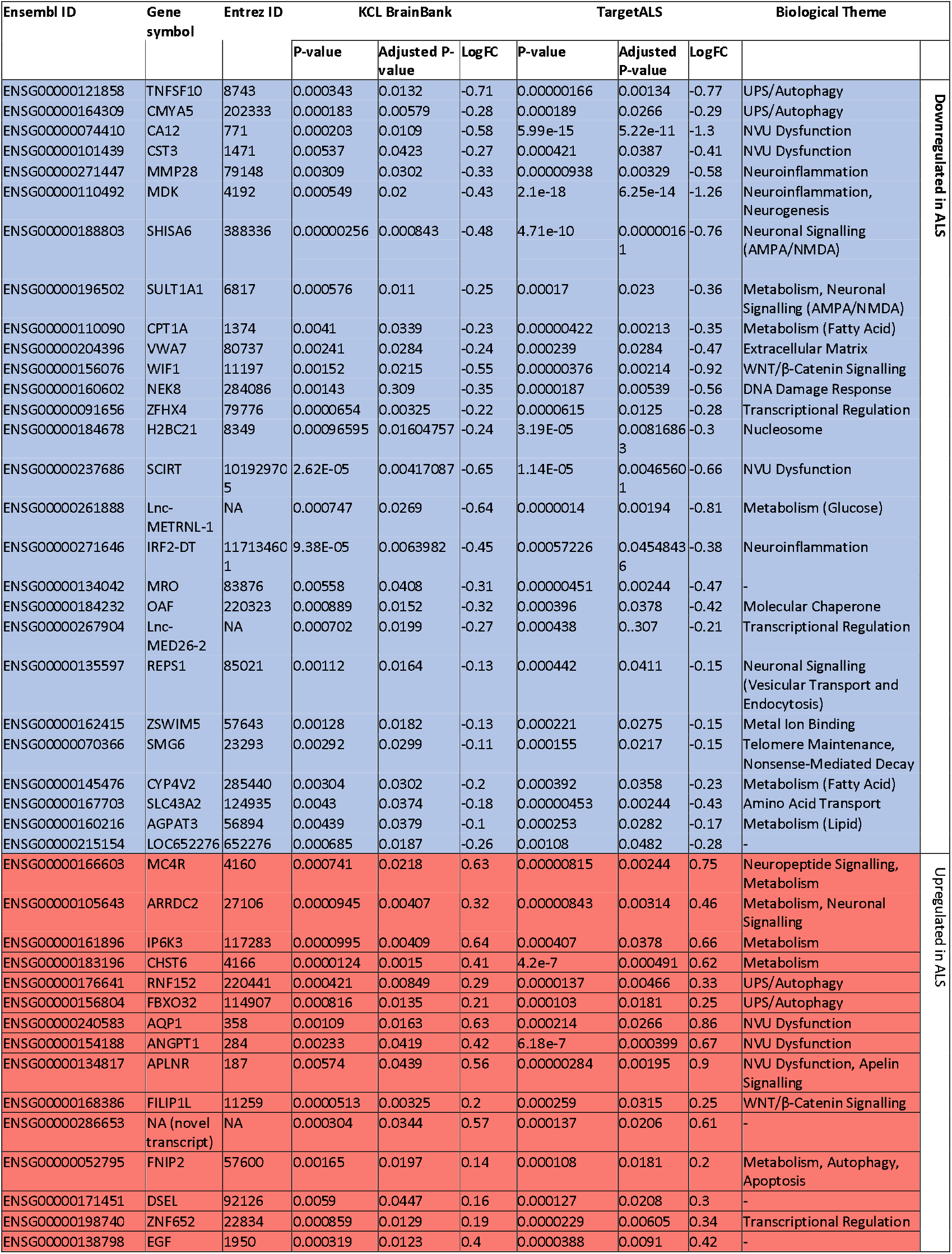
Genes significantly differentially expressed in both datasets. Red: Upregulated; Blue: Downregulated’ LogFC: Log2 of Fold Change

### Enrichment Analyses

To compensate for the different numbers of differentially expressed genes in the two datasets when using 0.05 as the corrected p-value threshold (2,290 genes in KCL BrainBank and 402 in TargetALS), we applied an additional threshold of logFC > 0.2 to the KCL BrainBank dataset. This yielded differential expression for 1,013 genes, which were uploaded to MetaCore for enrichment analysis along with the 402 differentially expressed genes in TargetALS. We performed analyses using MetaCore Pathway Maps, Process Networks, Disease Networks and GO processes. *Supplementary Tables 2* and *3* list all enrichments with their *p*-value, false FDR adjusted *p*-value, and the genes from each dataset contributing to each enrichment.

#### Process Networks: neuropeptide signalling is altered in ALS in both datasets

Figures *2A* and *2B* display the most significant MetaCore Process Networks identified in the two datasets. Among the most significant processes in KCL BrainBank are several neurological-related processes (*Transmission of nerve impulse, Neuropeptide signalling pathways* and *Development Neurogenesis Axonal guidance*) and processes related to muscle contraction. The most significant process network in TargetALS is *Signal Transduction Neuropeptide signalling pathways*, which was the third most significant KCL BrainBank process network. In TargetALS, among the most enriched networks (albeit not significant) are several processes related to inflammation (*interferon signalling* and *NK cell cytotoxicity*).

**Figure 2.**
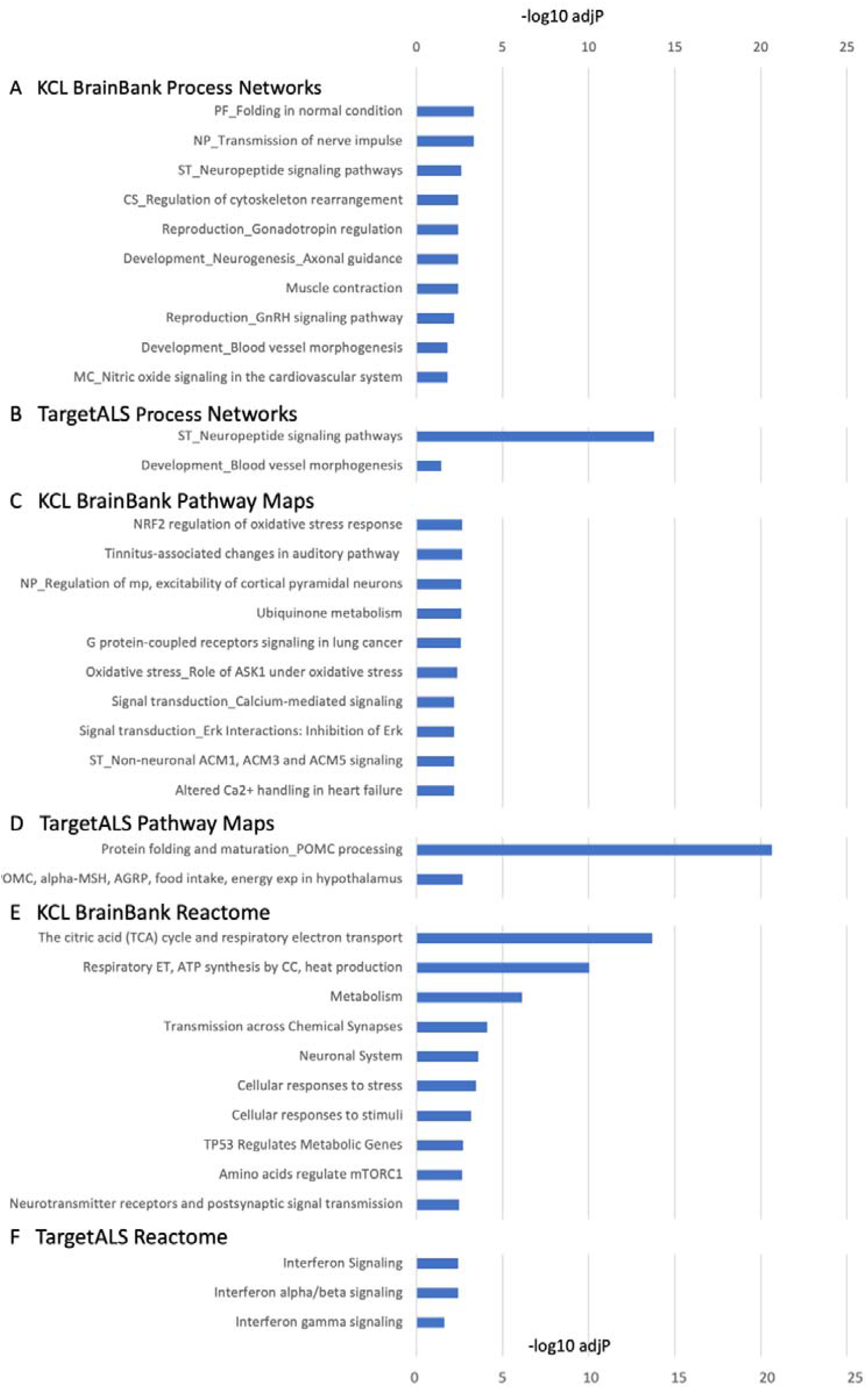
Enrichment analysis results in the two datasets. The ten most significant enrichments are shown, sorted by adjusted p-value, or fewer if there are less than ten significant after multiple testing correction. A) KCL BrainBank MetaCore Process Networks; B) TargetALS MetaCore Process Networks; C) KCL BrainBank MetaCore Pathway Maps; D) TargetALS MetaCore Pathway Maps; E) KCL BrainBank Reactome; For clarity, in panel E, one of the two redundant enrichments was omitted (*Respiratory electron transport*, which is more generalised of the one included, *Respiratory electron transport, ATP synthesis by chemiosmotic coupling, and heat production by uncoupling proteins*). F) TargetALS Reactome. Following pathway abbreviations/names have been introduced: PF: Protein Folding; NP: Neurophysiological process; ST: Signal transduction; CS: Cytoskeleton; MC: Muscle contraction; POMC, alpha-MSH, AGRP, food intake, energy exp in hypothalamus: POMC, alpha-MSH and AGRP in regulation of food intake and energy expenditure in obesity in hypothalamus; Regulation of mp, excitability of cortical pyramidal neurons: Regulation of intrinsic membrane properties and excitability of cortical pyramidal neurons; Respiratory ET, ATP synthesis by CC, heat production: Respiratory electron transport, ATP synthesis by chemiosmotic coupling, and heat production by uncoupling proteins.

#### Pathway Maps: KCL BrainBank is enriched for neurophysiological processes, oxidative stress and signal transduction, Target ALS is for POMC processing

Figures *2C* and *2D* present the most significantly enriched pathway maps in the two datasets. Prominent themes in the KCL BrainBank enrichments were neurophysiological processes, oxidative stress and signal transduction. There were three significant TargetALS pathway maps; two related to pro-opiomelanocortin (POMC) processing and one to immune response. Interestingly, the most significant network in TargetALS, POMC processing (Figure *3*), includes several agonists of *MC4R* which is significantly upregulated in both datasets.

**Figure 3.**
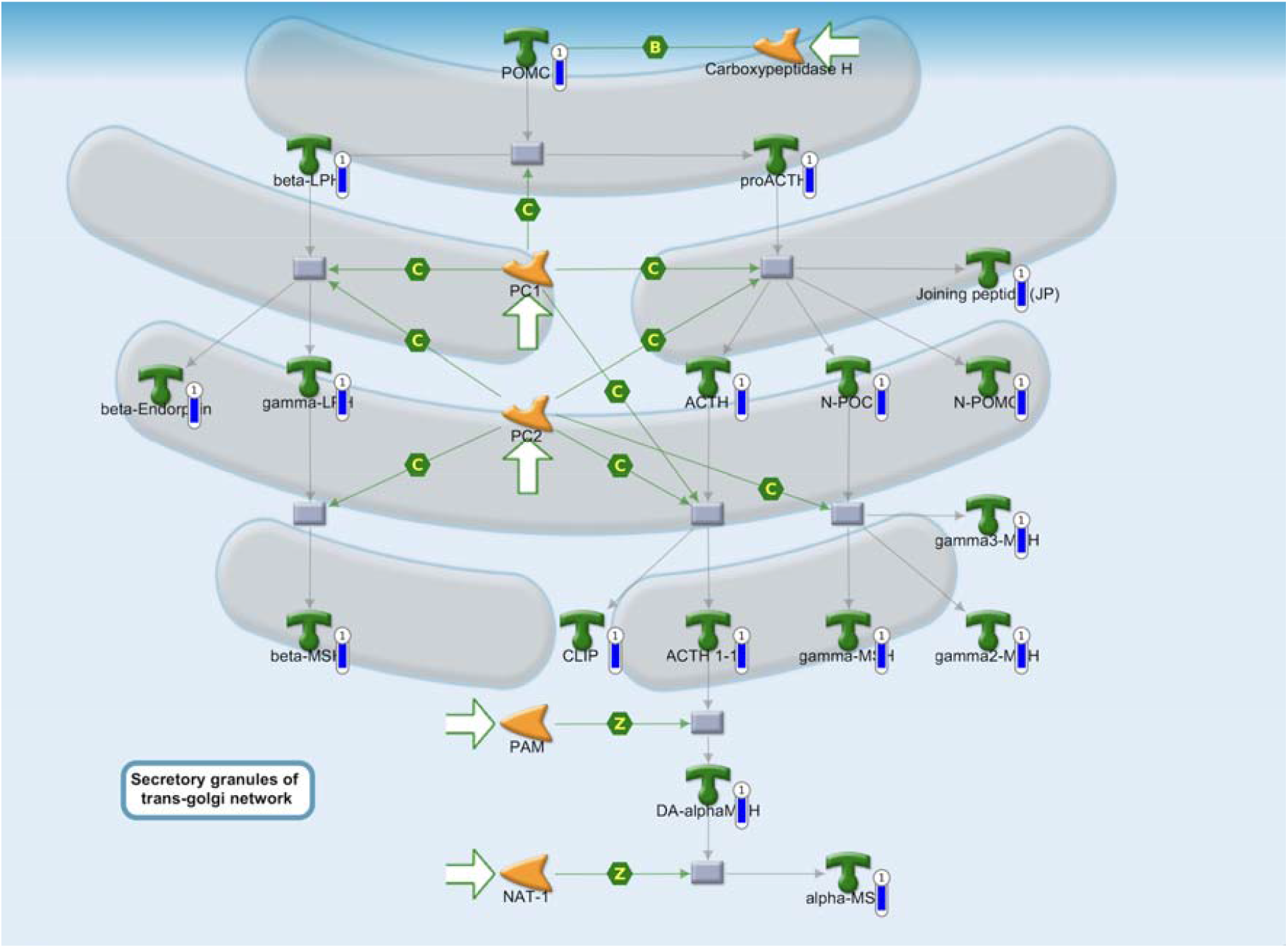
Protein folding and maturation POMC processing pathway map from MetaCore. Blue barometers represent downregulated genes from the TargetALS dataset. The descriptions of what the other symbols represent are available in *Supplementary Figure 1*.

#### KCL BrainBank is enriched for nervous system-specific processes while TargetALS is for immunological processes

Enrichments for MetaCore *Diseases by Biomarkers*, GO Biological Processes and Reactome pathways are concordant in the two datasets. The KCL dataset showed clear nervous system related enrichments. These included disease enrichments (Heredodegenerative diseases, Huntington’s disease, Central Nervous System diseases among others), GO Processes relevant to nervous system development, and Reactome pathways related to the functioning of the nervous system, such as the transmission of chemical synapses. TargetALS was enriched for immune system related diseases and processes, including diseases connected with response to various viral infections, GO Processes related to immune defence, and Reactome pathways pertaining to interferon signalling (Figures *2E* and *2F*).

### Correlation of neuropeptides and receptors expression with ALS phenotypic attributes

As neuropeptide signalling was the only altered biological network in both datasets, and neuropeptides have been implicated with the ALS clinical phenotype^14-16^, we decided to test the correlation between the expression of neuropeptides and their receptors with age at onset and disease duration of the ALS patients in both datasets. Several neuropeptides and receptors were found to be significantly correlated with age at onset and disease duration in at least one dataset (*Table 3*). *NPBWR1* was found to be significantly correlated with age at onset in both KCL BrainBank and TargetALS after multiple testing correction, whilst TAC3 was nominally significantly correlated with age at onset in TargetALS and after multiple testing correction in KCL BrainBank. *GNRH1* and *TACR1* were nominally significantly correlated with survival and *SSTR1* with age at onset in both datasets. Interestingly all neuropeptides and receptors that were significantly correlated with the age of onset of ALS in at least one dataset had a negative coefficient. *Figure 4A* shows *NPBWR1* expression against age of onset. Complete results can be found in *Supplementary Tables 4, 5, 6* and *7*.

**Table 3.**
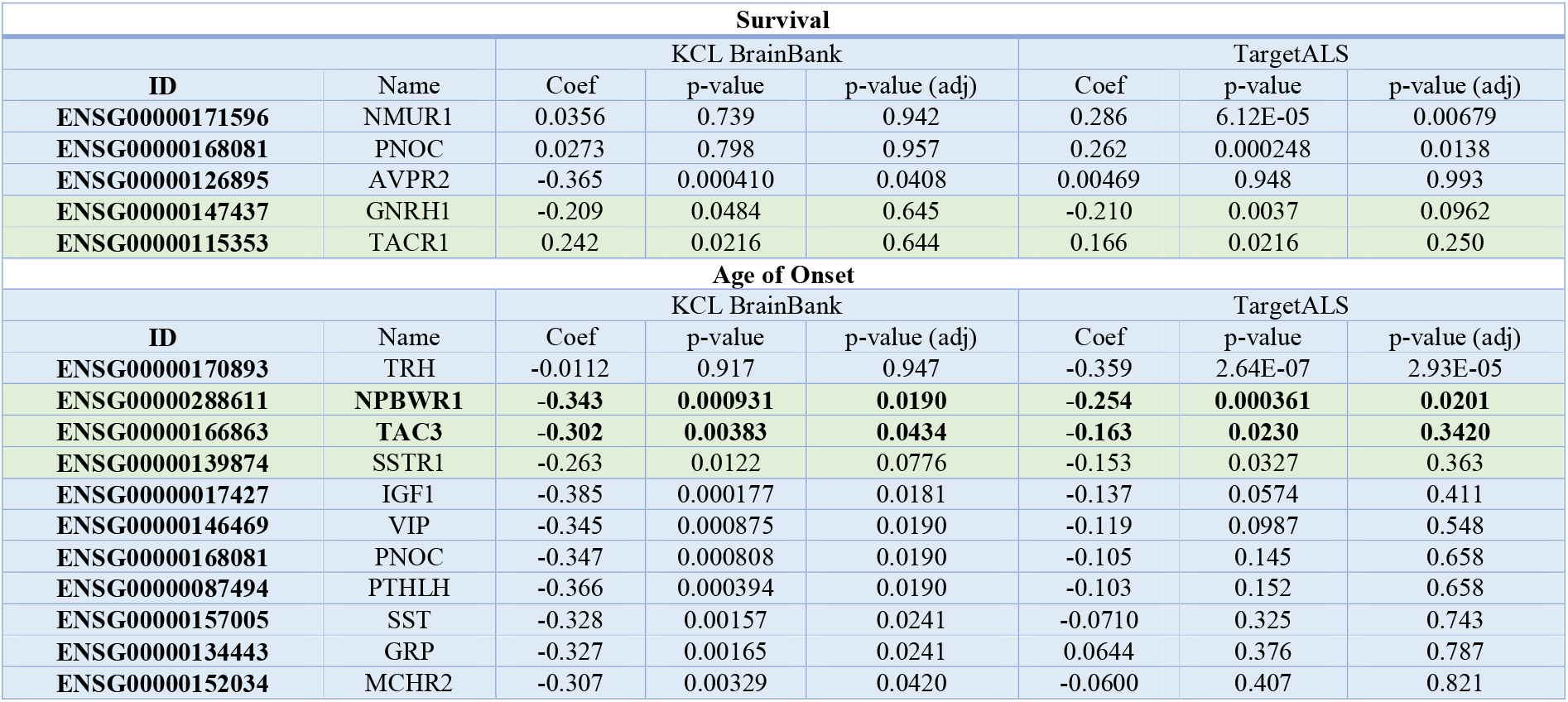
Results of the correlation analysis between neuropeptides and receptors, and clinical features. Only results that are significant after FDR correction in at least one dataset, or nominally significant in both are displayed. Correlations that are at least nominally significant in both datasets are coloured in green, and those that are also significant after FDR correction in one dataset are also in bold.

**Figure 4:**
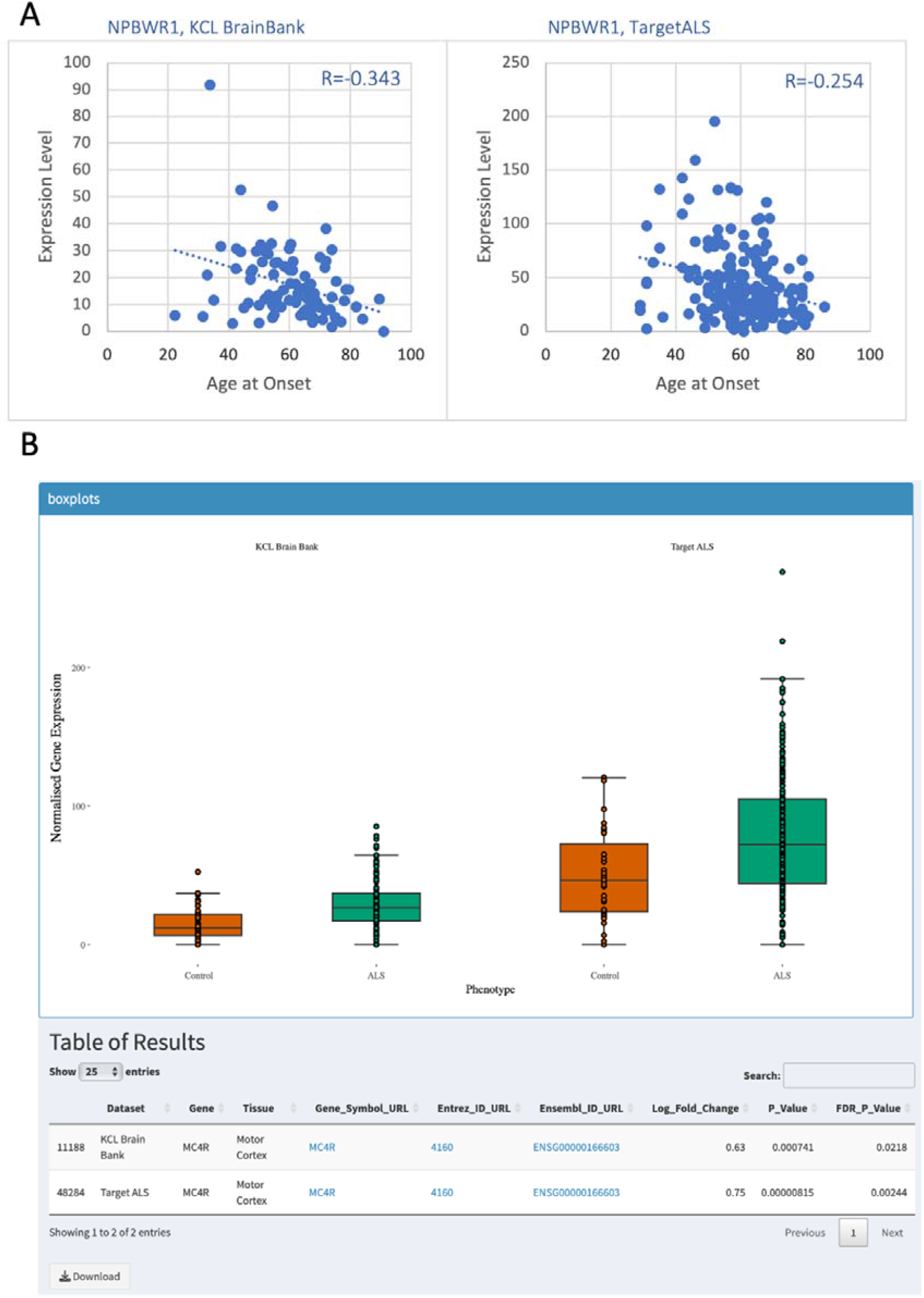
A) Correlation analysis between *NPBWR1* gene expression and age of onset in KCL BrainBank and TargetALS. B) Screenshot from ALS Gene Expression Explorer showing expression of *MC4R*, which was significantly upregulated in both datasets.

### Cell Type Composition

Cell type composition analysis is shown in *Figure 5*. There was a significantly smaller proportion of endothelial cells in cases compared to controls in both datasets (KCL BrainBank: F = 8.673, *p*-value = 0.00369; TargetALS: F = 6.423, *p*-value = 0.0119). In the KCL dataset, cases also displayed higher neuronal cell (F = 25.256, *p*-value = 1.28 × 10^−6^) and lower oligodendrocyte levels (F = 13.633, *p*-value = 0.000301) whereas, in TargetALS, astrocytes were more prevalent amongst cases (F = 5.466, *p*-value = 0.0203). The full results are available in *Supplementary Table 8*.

**Figure 5.**
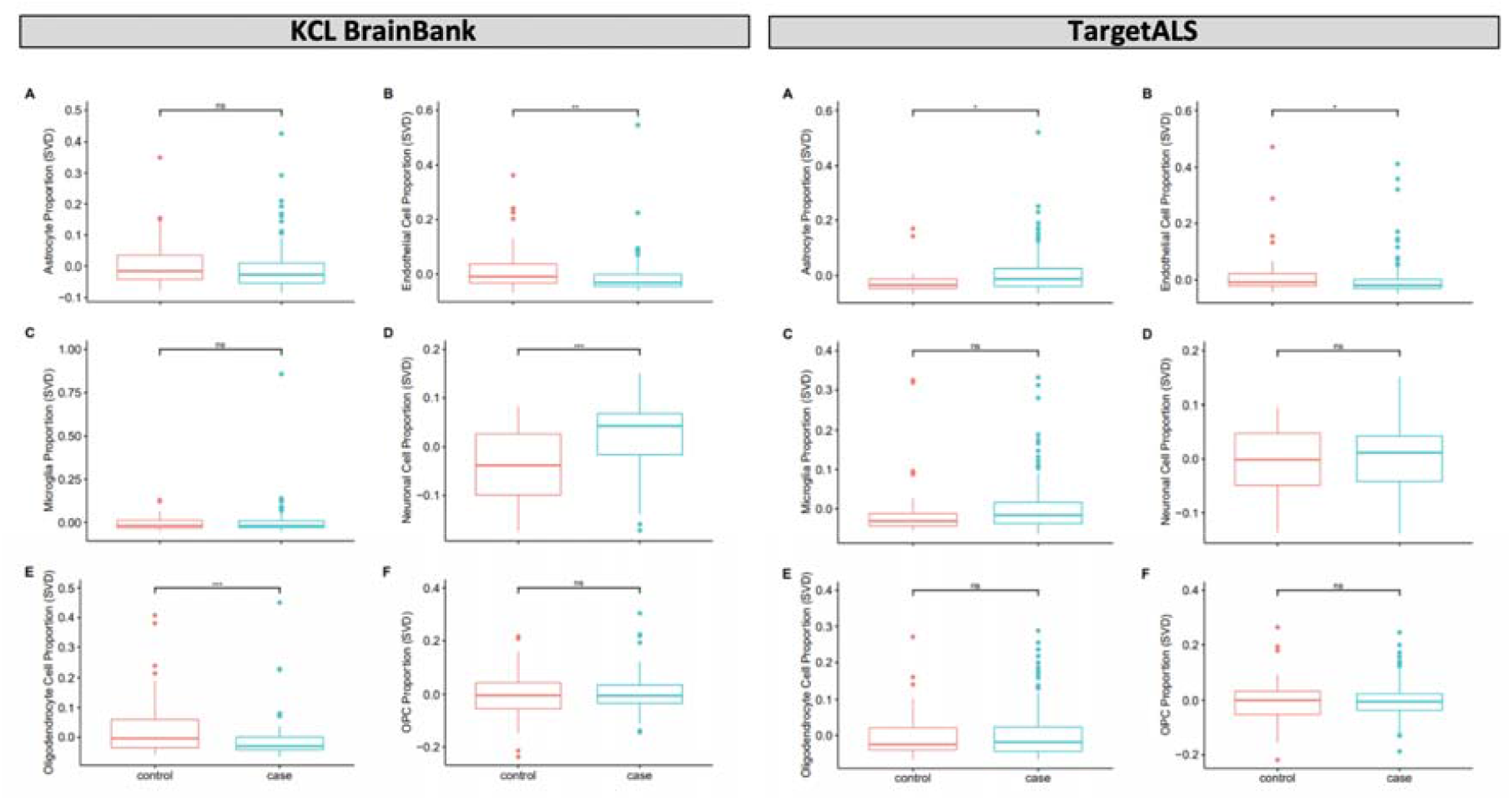
Case-control cell composition analysis in KCL BrainBank (left panel) and TargetALS (right panel), using singular decomposition values generated by the BRETIGEA program for six cell types: A) Astrocytes, B) Endothelial Cells, C) Microglia, D) Neurons, E) Oligodendrocytes, F) Oligodendrocyte Progenitor Cells. Significance was assessed using one-way ANOVA. ns = non-significant, ns = non-significant, * < 0.05, ** < 0.01, *** < 0.001. Results were corrected for age and sex. Significance was assessed using one-way ANOVA. Results were corrected for age and sex. Controls are in red and cases in green.

#### ALS Gene Expression Explorer

The ALS Gene Expression Explorer web application (https://alsgeexplorer.er.kcl.ac.uk) enables users to explore and compare the expression levels for specific genes in ALS cases and controls in the two datasets. *Figure 4B* displays an example gene (*MC4R*) which is differentially expressed in both datasets.

## Discussion

By leveraging motor cortex expression data from two independent large datasets (a total of 224 ALS cases and 79 controls), we reported ALS gene expression profiles that indicate the involvement of various biological processes in the pathology of ALS. We also identified genes in the neuropeptide signalling pathway that showed potential as biomarkers of ALS onset and progression. Differential expression analysis identified 2,290 and 402 differentially expressed genes in KCL BrainBank and TargetALS cases, respectively. Synapse-related processes, such as GABAergic neurotransmission, calcium-mediated signalling and postsynaptic signalling were significantly enriched in KCL BrainBank, whilst TargetALS carried an immune-system related signature, with strong enrichments for interferon signalling and NK cell neurotoxicity. This is most likely a product of the molecular heterogeneity of ALS, as these two profiles reflect previously hypothesised mechanisms of ALS pathogenesis and correspond to the predominant cell types identified in these datasets (neuronal cells in KCL BrainBank and astrocytes in TargetALS). Changes in the excitability patterns of GABAergic interneurons influence alterations in motor cortex circuitry, which has been proposed as a possible trigger for the spread of ALS pathology^17,18^. Previous work has also implicated NK cells, which secrete interferon-γ and are upregulated in the motor cortex and spinal cord of ALS patients^19,20^ as being modulators of motor neuron degeneration^21^, with interferon signalling pathways being upregulated in spinal cord astrocytes in a *SOD1* ALS mouse model^22^.

Despite the diversity in the KCL BrainBank and TargetALS gene expression profiles, the neuropeptide signalling pathway was enriched in both datasets, which suggests that this is a key feature of ALS pathogenesis. The KCL BrainBank and TargetALS datasets exhibited both unique and shared differential expression of several neuropeptide ligands and receptors. One gene, melanocortin receptor 4 (*MC4R*), was significantly upregulated in both datasets. The role of the melanocortin system in ALS has largely been overlooked, despite *MC4R* activation inducing hypermetabolism, increased energy expenditure and cachexia; three processes associated with increased lower motor neuron weakness and poor prognosis in ALS patients^23-27^. Furthermore, *MC4R*-mediated signalling in the motor cortex also enhances the activity of sympathetic preganglionic neurons in the spinal cord, which modulates skeletal muscle contraction^28,29^, and rare (MAF <0.01) disruptive and damaging missense variants in *MC4R* might increase the risk of ALS (Beta = 2.15 SE = 1.52, *p*-value = 0.037, http://databrowser.projectmine.com). Therefore, we suggest further investigation on *MC4R* as a potential candidate target gene for future biomarker and screening studies.

Several *MC4R* agonists are involved in the trans-Golgi network protein folding and maturation pathway, which was the most significantly downregulated pathway in TargetALS. This pathway is controlled by the *MC4R* ligand pro-opiomelanocortin (*POMC*), which acts as a precursor peptide for the *MC4R* agonists^30^. Hence, aberrant protein degradation linked to both neuropeptide signalling dysregulation and endoplasmic reticulum associated stress could be a triggering event for ALS pathogenesis. We observed reduced expression of *POMC* in TargetALS (LogFC = -1.12, adjusted *p*-value = 3.6E-05), which would undoubtedly affect the processing of several peptides reliant on *POMC* cleavage in this pathway (*β-endorphin, ⍰-MSH, β-MSH*). Their expression is linked to anti-inflammatory processes in neurodegenerative diseases. For instance, *β-endorphin*, which normally reduces the production of pro-inflammatory cytokines such as *interferon-γ, IL-1β* and *IL-2*^31^ is found at lower levels in the cerebrospinal fluid of Alzheimer’s disease patients^32^. Similarly, both *⍰-MSH* and *β-MSH* are cytokine antagonists that protect against excitotoxic and apoptotic signalling effects in Parkinson’s and Alzheimer’s disease models^33,34^; actions that are also perturbed in ALS as evidenced by similarly enriched pathways in this study. Therefore, the effects that *MC4R*-*POMC* signalling has on inflammatory processes could also explain the significant enrichment of immune-related processes observed in the TargetALS dataset.

To further investigate how neuropeptide signalling affects clinical phenotype, we performed a correlation analysis of neuropeptides and their receptors (102 neurotransmitters and attributes present in KCL BrainBank and 111 in TargetALS) with clinical attributes. Several neuropeptides and receptors significantly correlated with age of onset and survival in both datasets. *NPBWR1* was significantly correlated with the age of onset after multiple testing correction in both datasets (KCL BrainBank r = -0.34, adjusted *p*-value = 0.019; TargetALS r = -0.25, adjusted *p*-value = 0.02). Interestingly, *NPBWR1* is expressed by GABAergic interneurons and when activated, can dampen their inhibitory effects^35^. Therefore, increased *NPBWR1* expression could have a greater impact on the aforementioned alterations in motor cortex circuitry, which would corroborate with the earlier age of onset in patients with higher *NPBWR1* levels. Likewise, expression of the somatostatin receptor *SSTR1*, which also resides on inhibitory GABAergic interneurons^36^, was nominally negatively correlated with age of onset in the KCL BrainBank. We also found a nominally significant correlation between *TACR1* and *GNRH1* levels and survival in both datasets. Both *TACR1* and *GNRH1* can stimulate the production and release of arachidonic acid^37,38^, a polyunsaturated fatty acid whose levels can distinguish ALS patients from mimics and controls and positively correlates with disease progression^39^, with higher levels also found in spinal cord motor neuron cultures^40^. Therefore, the role of the neuropeptide signalling system on lipid metabolism and cortical signalling in ALS is warranted to assess its full impact on clinical phenotype.

When reviewing the 44 differentially expressed genes which were shared in both datasets, we found that many were involved in integral themes of ALS pathogenesis, such as carbohydrate/lipid metabolism, mitochondrial energy homeostasis (*SULT1A1, IP6K3, CPT1A*)^41-43^, and neurovascular unit dysfunction (*AQP1, APLNR, CA12, AGPT1, CST3*)^44-48^. Further investigations of these genes are needed to determine if they are the drivers of alterations in these pathways in ALS.

Our study has some limitations which are primarily linked to the fact that we identified gene expression signatures in post-mortem tissue. Firstly, as the cells we derive expression for have survived the neurodegenerative processes of ALS, we cannot infer if the changes we are observing are causing or are a consequence of the ALS phenotype. We do not have information on the staging of ALS in these patients so cannot attribute the expression profiles to a certain point of ALS progression. Furthermore, there were 5.7 times more differentially expressed genes in KCL BrainBank than in TargetALS. Although this might be linked to the larger number of controls in the KCL datasets (∼3 times more than in TargetALS), there could also be several external factors influencing these datasets, such as library preparation, RNA extraction, sample quality, and population variability in gene expression.

In summary, we identified several key modulatory themes of ALS pathogenesis as well as identified genes in the neuropeptide signalling pathway that showed potential as diagnostic and prognostic biomarkers. There is a great need for accurate biomarkers in ALS, particularly for biomarkers that predict the age of onset. These could be explored to inform the study of the pre-symptomatic stages of ALS and support the development of pre-symptomatic diagnostic approaches^49^.

## Data Availability

KCL BrainBank RNAseq data is available, on reasonable request, from the corresponding author. All raw RNAseq data from TargetALS samples can be requested by emailing.
Results for individual genes can be visually explored at https://alsgeexplorer.er.kcl.ac.uk/. All data analysis scripts used in this study are available at https://github.com/rkabiljo/RNASeq_Genes_ERVs and https://github.com/rkabiljo/DifferentialExpression_Genes.

https://alsgeexplorer.er.kcl.ac.uk/

https://github.com/rkabiljo/RNASeq_Genes_ERVs

https://github.com/rkabiljo/DifferentialExpression_Genes

## Ethics declaration

*Post-mortem* tissue samples from King’s College London were collected under the ethical approval of the MRC London Neurodegenerative Diseases Brain Bank and under the regulations of the Human Tissue Act UK 2014. All post-mortem tissue was donated to the MRC London Neurodegenerative Diseases Brain Bank under standard ethical and Human Tissue Act procedures, with informed consent provided by the next of kin. Data generated from this material were anonymized and analysed on a high-performance computing cloud (https://www.maudsleybrc.nihr.ac.uk/facilities/rosalind/) with data protection protocols in accordance with Department of Health Policy (UK) and the security standards set by the National Data Guardian. Ethical approval to process and analyse *post-mortem* samples stored at King’s College London was provided by a local ethics committee at the Institute of Psychiatry, Psychology & Neuroscience, King’s College London, and the MRC London Neurodegenerative Diseases Brain Bank.

## Data availability

KCL BrainBank RNAseq data is available, on reasonable request, from the corresponding author. All raw RNAseq data from TargetALS samples can be requested by emailing. Results for individual genes can be visually explored at https://alsgeexplorer.er.kcl.ac.uk/. All data analysis scripts used in this study are available at https://github.com/rkabiljo/RNASeq_Genes_ERVs and https://github.com/rkabiljo/DifferentialExpression_Genes.

## Acknowledgements

The TargetALS Human Postmortem Tissue Core, New York Genome Center for Genomics of Neurodegenerative Disease, the Amyotrophic Lateral Sclerosis Association and the Tow Foundation.

Human post-mortem tissue was obtained from the Medical Research Council London Neurodegenerative Diseases Brain Bank (King’s College London). The London Neurodegenerative Diseases Brain Bank at KCL has received funding from the MRC and through the Brains for Dementia Research project (jointly funded by Alzheimer’s Society and Alzheimer’s Research UK)’.

This work was carried out on King’s College HPC Create: King’s College London. (2022). King’s Computational Research, Engineering and Technology Environment (CREATE). Retrieved March 2, 2022, from https://doi.org/10.18742/rnvf-m076, and on the Pawsey Supercomputing Research Centre with funding from the Australian Government and the Government of Western Australia.

RK is funded by MND Scotland, grant number RE14937.

HM is funded by GlaxoSmithKline and the KCL funded centre for Doctoral Training (CDT) in Data-Driven Health.

GH is funded by Perron Institute for Neurological and Translational Science and the KCL funded centre for Doctoral Training (CDT) in Data-Driven Health.RJBD is supported by the following: (1) NIHR Biomedical Research Centre at South London and Maudsley NHS Foundation Trust and King’s College London, London, UK; (2) Health Data Research UK, which is funded by the UK Medical Research Council, Engineering and Physical Sciences Research Council, Economic and Social Research Council, Department of Health and Social Care (England), Chief Scientist Office of the Scottish Government Health and Social Care Directorates, Health and Social Care Research and Development Division (Welsh Government), Public Health Agency (Northern Ireland), British Heart Foundation and Wellcome Trust; (3) The BigData@Heart Consortium, funded by the Innovative Medicines Initiative-2 Joint Undertaking under grant agreement No. 116074. This Joint Undertaking receives support from the European Union’s Horizon 2020 research and innovation programme and EFPIA; it is chaired by DE Grobbee and SD Anker, partnering with 20 academic and industry partners and ESC; (4) the National Institute for Health Research University College London Hospitals Biomedical Research Centre; (5) the National Institute for Health Research (NIHR) Biomedical Research Centre at South London and Maudsley NHS Foundation Trust and King’s College London; (6) the UK Research and Innovation London Medical Imaging & Artificial Intelligence Centre for Value Based Healthcare; (7) the National Institute for Health Research (NIHR) Applied Research Collaboration South London (NIHR ARC South London) at King’s College Hospital NHS Foundation Trust.

AAC is an NIHR Senior Investigator (NIHR202421). This research is part an EU Joint Programme - Neurodegenerative Disease Research (JPND) project. The project is supported through the following funding organisations under the aegis of JPND: Medical Research Council; Economic and Social Research Council and the Motor Neurone Disease Association. Funding for open access charge: UKRI.

AI is funded by the Motor Neurone Disease Association. This study represents independent research partly funded by the National Institute for Health Research (NIHR) Biomedical Research Centre at South London and Maudsley NHS Foundation Trust and King’s College London. BA acknowledges funding through a Pre-doctoral Fellowship from the NIHR (NIHR301067). We acknowledge use of the research computing facility at King’s College London, Rosalind (https://rosalind.kcl.ac.uk), which is delivered in partnership with the National Institute for Health Research (NIHR) Biomedical Research Centres at South London & Maudsley and Guy’s & St. Thomas’ NHS Foundation Trusts and part-funded by capital equipment grants from the Maudsley Charity (award 980) and Guy’s and St Thomas’ Charity (TR130505). The views expressed are those of the author(s) and not necessarily those of the NHS, the NIHR, King’s College London, or the Department of Health and Social Care.

