## Supplementary materials for "Transcriptomics Analyses of ALS Post-mortem Motor Cortex highlight alteration and potential biomarkers in the Neuropeptide Signalling pathway"

### Supplementary Methods

### RNAseq protocol for samples from the MRC London Neurodegenerative Diseases Brain Bank (King's College London)

#### *Purification, isolation and quality control*

The  30mg tissue block for RNA was homogenised using a Qiagen PowerLyzer 24 Homogenizer. Total RNA was purified from the homogenate using the standard protocol of the RNeasy Lipid Tissue Mini Kit (Qiagen), with on-column DNAse digestion. RNA integrity was estimated using Agilent Bioanalyzer 2100's RNA 6000 Nano assays. RNA quantification was performed using a NanoDrop.

#### *Library preparation and RNA sequencing*

Library preparation was performed using the standard Illumina TruSeq Stranded Total RNA Sample Preparation Guide with Ribo-Zero Human/Mouse/Rat (October 2013 Rev E.). Fragmentation steps were tailored to degrees of degraded RNA samples using Agilent Bioanalyzer 2100's Nano assay results from the previous section. Libraries were validated using an Agilent Bioanalyzer 2100 to assess fragment size distribution. Library concentrations were estimated using a Qubit RNA High Sensitivity Assay Kit. Nanomolar (nM) concentrations were estimated using nM = ng/ul X (1500/Average bp). Libraries were sequenced using Illumina HiSeq 4000 flow cells with 150bp paired-end reads with a target depth of 30 million clusters (60 million reads per sample).

### TargetALS Sample Selection

In this study, we included 234 samples from 132 subjects from the TargetALS dataset. The distribution of the samples was 112 samples in the lateral motor cortex, 112 in the medial motor cortex and a further 10 samples in unspecified motor cortex. 30 subjects contributed a single sample, while 102 subjects contributed two samples from two different regions of the motor cortex. Figure S1 shows the PCA of the 234 samples, coloured by the motor cortex region.

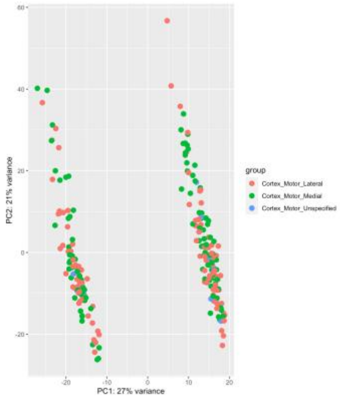


**Figure S1**. PCA of 241 selected samples from TargetALS. Coloured by motor cortex region.

To assess whether merging the three motor cortex regions to gain statistical power was a suitable approach, we performed separate case-control differential expression analyses of the lateral motor cortex and medial motor cortex and compared them to the results of the same analysis when all motor cortex regions are pooled together. As the unspecified motor cortex includes only ten samples, we did not perform a separate differential expression analysis. In the lateral motor cortex (96 ALS cases and 16 controls), 8 genes crossed genome wide significance (corrected p-value<0.05). All these genes are significant in the merged dataset. In the medial motor cortex (93 ALS cases and 19 controls), 42 genes crossed genome wide significance (corrected p-value<0.05), with 39 of these genes (93%) also being significant in the merged dataset. As the merged dataset gives us more statistical power and still detects the vast majority of differentially expressed genes in the two smaller subsets, we decided to use the merged dataset in our study.

### Post-sequencing Bioinformatics Analyses

Clip adapter sequences and low-quality reads were removed using BBDuk (http://jgi.doe.gov/data-and-tools/bb-tools/). We used STAR v2.7.10a[1] and Homo_sapiens.GRCh38.104.gtf reference assemblies for alignment of RNAseq reads and HTSeq[9] version 2.0.0 for transcript quantification. This pipeline is available at https://github.com/rkabiljo/RNASeq_Genes_ERVs . For the KCL BrainBank dataset, we included age, sex, RIN number, PMD and a surrogate variable obtained by SVA[2] covariates. In addition to these, we also included the sequencing centre as a covariate in the TargetALS dataset. For differential expression analysis, we used DESeq2, with the Wald test used for differential expression, and the Benjamini-Hochberg method used for *p*-value adjustment. The differential expression code is available at https://github.com/rkabiljo/DifferentialExpression_Genes. For MA Plots, apeglm was used for LFC shrinkage[3]. For enrichment analysis we used MetaCore. Due to a non-one-to-one mapping of the MetaCore algorithm of uploaded gene lists to their proprietary concept of network objects, the resulting enrichment lists consisted of 1,004 objects for KCL BrainBank and 366 for TargetALS. For background genes, we used a list of 41,709 transcripts which had at least 10 counts in at least 5 subjects, which were subsequently mapped in MetaCore to 26,484 network objects. To assess if gene enrichment results were a product of the contribution of different cell types in the bulk RNA sequencing data, we used the BRETIGEA cell type estimation R package (version 1.0.3) under default settings to derive cell proportions for the following cell types; astrocytes, endothelial cells, microglia, neurons, oligodendrocytes and oligodendrocyte progenitor cells (OPCs). Case-control differences in cell type proportions were assessed using one-way ANOVA corrected for age and sex, with a p-value of $< 0.05$ denoting significance. Pearson correlation between melanocortin receptors and their ligands, POMC and AGRP, was performed using the *cor* function of WGCNA[4] (version 1.71).

ALS Gene Expression Explorer Webserver

The webserver code is publicly available on GitHub and requires R version 4.1.2 ≤.
